# Deep Learning-Based Detection of Focal Cortical Dysplasia in Children: External Validation of the MELD Graph and 3D-nnUNet pipelines

**DOI:** 10.64898/2026.04.21.26351368

**Authors:** Andrea Dell’Orco, Enrico De Vita, Felice D’Arco, Annalena Lange, Theodor Rüber, Angela M. Kaindl, Mike P. Wattjes, Ulrich-Wilhelm Thomale, Lena-Luise Becker, Anna Tietze

## Abstract

Focal cortical dysplasias (FCDs) are one of the most common structural causes of drug-resistant epilepsy in children but are frequently subtle and difficult to detect on conventional MRI. Many automated lesion detection methods have therefore been proposed to support neuroradiological assessment. In this study, we externally validated two recently developed deep-learning approaches for FCD detection, MELD Graph and 3D-nnUNet, in a pediatric cohort.

In this retrospective single-center study, brain MRI scans of 71 children evaluated for epilepsy were analyzed, including 35 MRI-positive patients with suspected FCD and 36 MRI-negative cases based on the primary radiology reports. Both models were applied to standard 3D T1-weighted and 3D FLAIR images. Detected lesions were reviewed by an experienced pediatric neuroradiologist and classified as true positive, false positive, or false negative. Clinical semiology and EEG findings were additionally evaluated for cases with false-positive detections.

At the lesion level, MELD Graph achieved a precision of 0.85 and recall of 0.52, while 3D-nnUNet achieved a precision of 0.91 and recall of 0.48. In the MRI-negative patients, MELD Graph produced more false-positive detections than 3D-nnUNet (0.53 vs. 0.14 false-positive lesions per patient). At the patient level, MELD Graph showed slightly higher sensitivity than 3D-nnUNet (0.63 vs. 0.54), whereas 3D-nnUNet demonstrated markedly higher specificity (0.86 vs. 0.56). Improved FLAIR image quality was associated with trends toward improved model performance.

Both models demonstrated high precision but moderate sensitivity, indicating that they are valuable decision-support tools but cannot replace expert neuroradiological evaluation. Optimized MRI acquisition protocols are needed to further improve automated lesion detection in pediatric epilepsy.

## Introduction

Epilepsy remains a major cause of morbidity and mortality in childhood [1,2]. About 25% of cases have structural causes, including perinatal or acquired brain injury, low-grade tumors, or vascular malformations, while around 5% are related to malformations of cortical development [2]. Among these, focal cortical dysplasias (FCDs) are the most frequent [3]. FCDs result from disordered neuronal maturation and cortical organization, are highly epileptogenic, and are histologically classified into three groups, with FCD type II being the most common subtype [3]. In a large study by Blümcke et al., specimens from more than 2,600 children undergoing epilepsy surgery were histopathologically analyzed, and approximately 28% were found to have FCDs [3]. However, FCDs are often subtle and can be difficult to detect on conventional MRI. This underscores the importance of evaluating lesion detection methods specifically in the pediatric population; however, such studies remain scarce [4].

Antiseizure medication (ASM) is the first-line treatment, but about one third of children develop drug-resistant epilepsy, meaning they continue to have seizures despite adequate trials of at least two ASM [5]. Currently, epilepsy surgery is the only treatment with the potential to achieve complete seizure freedom and eventually allow ASM discontinuation. Identifying the epileptogenic lesion is therefore a crucial element to determine whether surgery is a viable option.

Multimodal approaches have been proposed to improve diagnostic accuracy, including arterial spin labeling [6], edge-enhanced MP2RAGE (Magnetization Prepared Rapid Acquisition Gradient Echo with two inversion times) [7], positron emission tomography (PET)-based techniques, and single-photon emission computed tomography (SPECT) [8]. In addition, advanced post-processing of MRI data has become an important and evolving strategy to detect subtle abnormalities and reduce the number of MRI-negative cases. Different approaches have been described including voxel-based morphometry[9], voxel-based intensity analysis [10], and sulcal morphometry [11,12].

Automated FCD detection is attractive because it is relatively easy to implement in clinical practice and typically relies on standard MRI sequences such as 3D T1-weighted and FLAIR images. Since the manual detection can be challenging and time-consuming, these tools offer a valuable aid in the daily radiological workflow. Sensitivity and specificity must, however, be carefully balanced: missing a lesion is unacceptable, yet low specificity with excessive false positives increases the number of findings that require manual verification, ultimately reducing radiologist efficiency. Post-processing methods have been validated against human readers, with mixed results [13–15].

Addressing this challenge, the multi-center lesion detection (MELD) consortium has recently refined its machine-learning–based approach by incorporating graph neural networks (MELD Graph [16]), demonstrating significantly improved positive predictive values (PPV) in a large cohort of more than 500 patients. Moreover, a recent study [14] suggested voxel-wise convolutional neural network–based segmentation using nnU-Net architectures (hereafter referred to as 3D-nnUNet). Both methods are promising because they lead to a high detection rate while maintaining a low number of false positive results.

Although both methods are evaluated on a multicenter cohort, the datasets consisted predominantly of adult patients. Each method has previously been evaluated on small external pediatric cohorts of 9-10 patients each [14,17]. Therefore, comprehensive external validation in a larger pediatric population remains essential to reliably assess their performance.

The aim of our study was to validate the MELD Graph and 3D-nnUNet approaches in a pediatric population by comparing children with FCD to those with MRI-negative epilepsy. We hypothesized that both methods would perform comparably to the original publications, with a number of false-positive findings acceptable for radiological workflow, and that their performance would depend on MRI data quality.

## Methods

### Dataset

The study was approved by the local Ethics Committee at Charité (EA2/295/25) and was conducted according to the guidelines of good clinical practice. Informed consent was waived in this retrospective study by the local Ethics Committee.

We searched our clinical picture archiving and communication system (PACS) retrospectively from July 2019 to March 2025 to identify MRI studies of patients evaluated for seizures or epilepsy. MRI were acquired on Skyra 3T scanner (Siemens Healthineers, Erlangen, Germany) with a dedicated 64-channel head coil and an MRI protocol including sagittal 3D T1w (MPRAGE) and 3D FLAIR. MRI acquisition parameters are reported in Table 1.

**Table 1:**
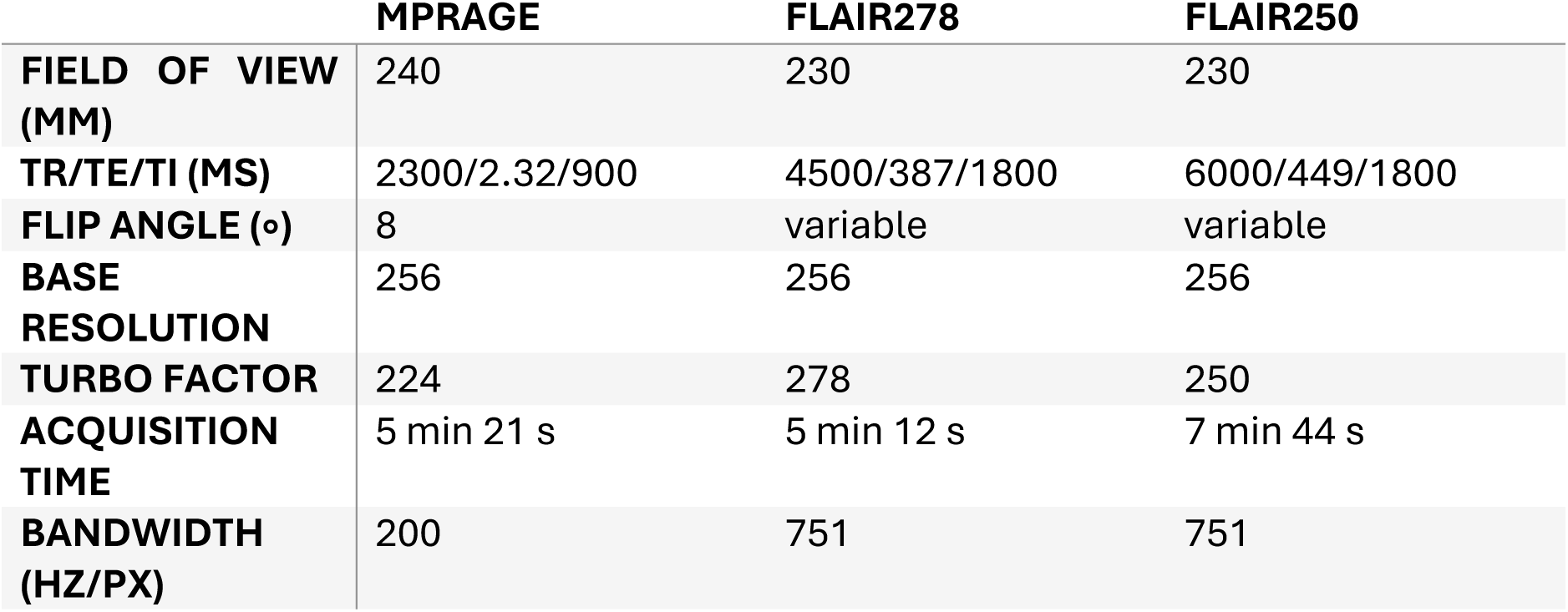
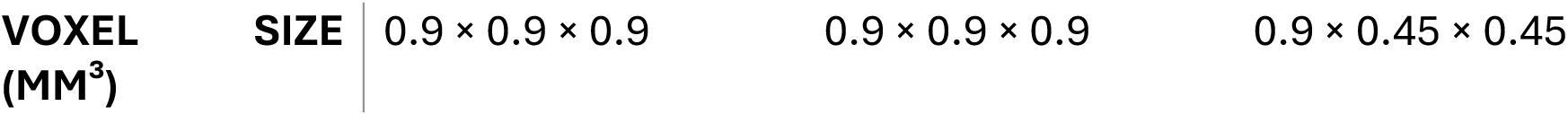
MRI acquisition parameters.

From 2024 onward, an improved FLAIR sequence was used, in which the turbo factor was decreased from 278 to 250 (hereafter referred to as FLAIR278 and FLAIR250, respectively) and reduced voxel-size. MRI parameters for the 3D T1w and the two versions of 3D FLAIR sequences are provided in Table 1. In addition, defaced example NIfTI files for the two sequences used in this study are available for download [18]. Only data free of motion artifacts or artifacts due to dental braces were included.

In addition to the MRI data, the original radiology report was retrieved. Studies were classified as MRI-positive (MRIpos) if a FCD had been identified, and as MRI-negative (MRIneg) if no lesion had been detected. Epilepsy patients with other structural abnormalities, including polymicrogyria, pachygyria, or epileptogenic tumors, were excluded from the analysis. Importantly, patients who were part of the original MELD training set were excluded.

### MELD Graph Segmentation

DICOM images exported from PACS were converted into NIfTI format with dcm2niix [19] and sorted in BIDS [20] format. A different harmonization model was used for each FLAIR version. After running the initial segmentation with FreeSurfer v7.2 [21], the harmonization step of the MELD Graph pipeline for all patients was run (*--harmo_only* flag), followed by the lesion prediction step.

Because incorrect FLAIR-to-T1w coregistration or poor FreeSurfer segmentation may affect lesion detection, the quality of coregistration, segmentation, and pial surface reconstruction was visually assessed for misregistration and missegmentation.

Quality control combined automated metrics obtained with the FSQC [22] tool, with subsequent visual inspection. FSQC, previously used in a similar study [23], has shown high agreement with expert ratings in a validation study [24]. Visual quality control was then performed independently by A.T. and A.D. following the MELD QC protocol [25]. Each subject was visually inspected and assigned a score of 1 (good segmentation), 2 (borderline), or 3 (to be excluded). Disagreements were resolved by consensus. Particular attention during visual review was given to subjects showing multiple outlying FSQC metrics based on z-scores of the individual metrics. Example images from the quality-control procedure are provided in Supporting Information, Fig. S2 and S3.

### 3D-nnUNet

To obtain the final predictions, we adopted the same ensemble strategy as described in the original publication [14]. FLAIR images were rigidly coregistered to the T1-weighted images using ANTs [26], using the default linear interpolation for the resampling. The coregistered T1-weighted and FLAIR images were then arranged into an nnU-Net-compatible dataset and segmented using the five independently trained models. The five model outputs were averaged voxel-wise, and the final segmentation was obtained by binarizing the resulting map at a threshold of 0.5, corresponding to a majority-vote rule across the ensemble.

### Neuroradiological and clinical evaluation of the detected lesions

The MELD Graph pipeline generates a PDF report highlighting each detected lesion together with a corresponding segmentation mask in NIfTI format. The 3D-nnUNet likewise produces lesion segmentation masks in NIfTI format. Lesions were annotated by anatomical location and hemisphere. MRI scans were initially assessed in routine clinical practice by the pediatric neuroradiologist on duty. All cases were subsequently re-evaluated by an experienced pediatric neuroradiologist (A.T.) with more than 10 years of experience in pediatric epilepsy imaging. This review compared the initial clinical reports with the MELD Graph and 3D-nnUNet segmentation masks, displayed as overlays on T1w and FLAIR images as well as all available MRI sequences in the PACS.

Lesions were classified as follows:

1. True positive lesions (TPL): identified by the neuroradiologist and detected by the model.
2. False positive lesions (FPL): detected by the model but not identified by the neuroradiologist as lesions, even after thorough re-evaluation.
3. False negative lesions (FNL): identified by the neuroradiologist but not detected by the model.
4. Retrospectively true positive lesions (rTPL), initially overlooked by the neuroradiologist but recognized on re-evaluation after review of the lesion-detection algorithm output. These retrospectively identified lesions were therefore reported separately and not incorporated into the predefined reference standard used for formal performance calculations.

Patient records with FPLs were re-evaluated by an experienced pediatrician specializing in pediatric neurology with extensive experience in pediatric epileptology. Clinical semiology and EEG findings were reviewed and compared with the results, with particular attention to whether the identified lesion was consistent with the clinical presentation. FPLs that were considered compatible with the clinical and/or EEG findings were subsequently reclassified as rTPL.

### Statistical evaluation

Statistical evaluation was performed separately for the two methods using the same set of metrics. Two complementary tasks were defined: a lesion-detection task and a patient-classification task.

The lesion-detection task evaluated the ability of the methods to localize individual lesions defined by neuroradiological assessment while minimizing FPLs that are time-consuming to review. In MRIpos patients, performance was quantified using lesion-level precision (also called positive predictive value, PPV) and recall (also called sensitivity). In MRIneg patients, no true lesions exist by definition; therefore, evaluation of the lesion-detection task was limited to quantifying FPL detections, summarized as the number of FPL per patient. Including MRIneg cases in lesion-level precision would be misleading, as they can contribute false positives but not true positives.

The patient-classification task was formulated as a binary decision problem, assessing whether a scan contained at least one lesion that was correctly detected by the model. Sensitivity and specificity were calculated based on the neuroradiological ground truth described above. At the patient level, true-positive patients (TPP) were defined as MRI-positive patients with at least one true lesion detected by the model; false-negative patients (FNP) as MRI-positive patients without any detected true lesion; false-positive patients (FPP) as MRI-negative patients with at least one incorrect detection; and true-negative patients (TNP) as MRI-negative patients without false-positive detections. Although recall and sensitivity are mathematically equivalent, we use the term recall for lesion-level detection and sensitivity for patient-level classification throughout the manuscript to avoid confusion between the two tasks.

Additionally, in order to investigate differences between models and FLAIR sequences, we fitted four regression models. For the lesion-detection task, we used a mixed-effects logistic regression with correctly detected lesion as the outcome (TPL = 1, FNL = 0), model as predictor, and FLAIR sequence, patient age, and sex as covariates. Subject was included as a random effect. For the patient-classification task, we used a logistic regression with correctly classified patient as the outcome and the same predictors and covariates.

To further assess the effect of FLAIR sequence on patient-level errors, we fitted two additional logistic regression models with false-positive patient status and false-negative patient status as outcomes, respectively. Both models included model, FLAIR sequence, and their interaction as predictors.

Formal definitions and formulas for all performance metrics are provided in Supporting Information S1.

## Results

### Dataset

The final dataset comprised 36 MRIneg and 35 MRIpos patients, with a total of 44 ground truth lesions. Among the MRIpos patients, 8 were histologically confirmed (3 FCD type 2A, 4 FCD type 2B, one FCD type 3A). In addition, one patient with tuberous sclerosis complex underwent resection for a tuber that is known to be very similar to FCD type 2A, two lesions turned out to be gangliogliomas, one lesion represented gliotic tissue, and one case remained histologically inconclusive. The remaining lesions were diagnosed based on imaging alone. Cohort demographics are summarized in Table 2. and Fig. 1. Of all patients, 44 patients (25 MRIpos, 19 MRIneg) were imaged using FLAIR278 and 27 using FLAIR250 (10 MRIpos, 17 MRIneg). A two-sided t-test revealed no significant age difference between participants scanned with FLAIR278 and those scanned with FLAIR250 (t = -0.79, p = 0.43).

**Figure 1:**
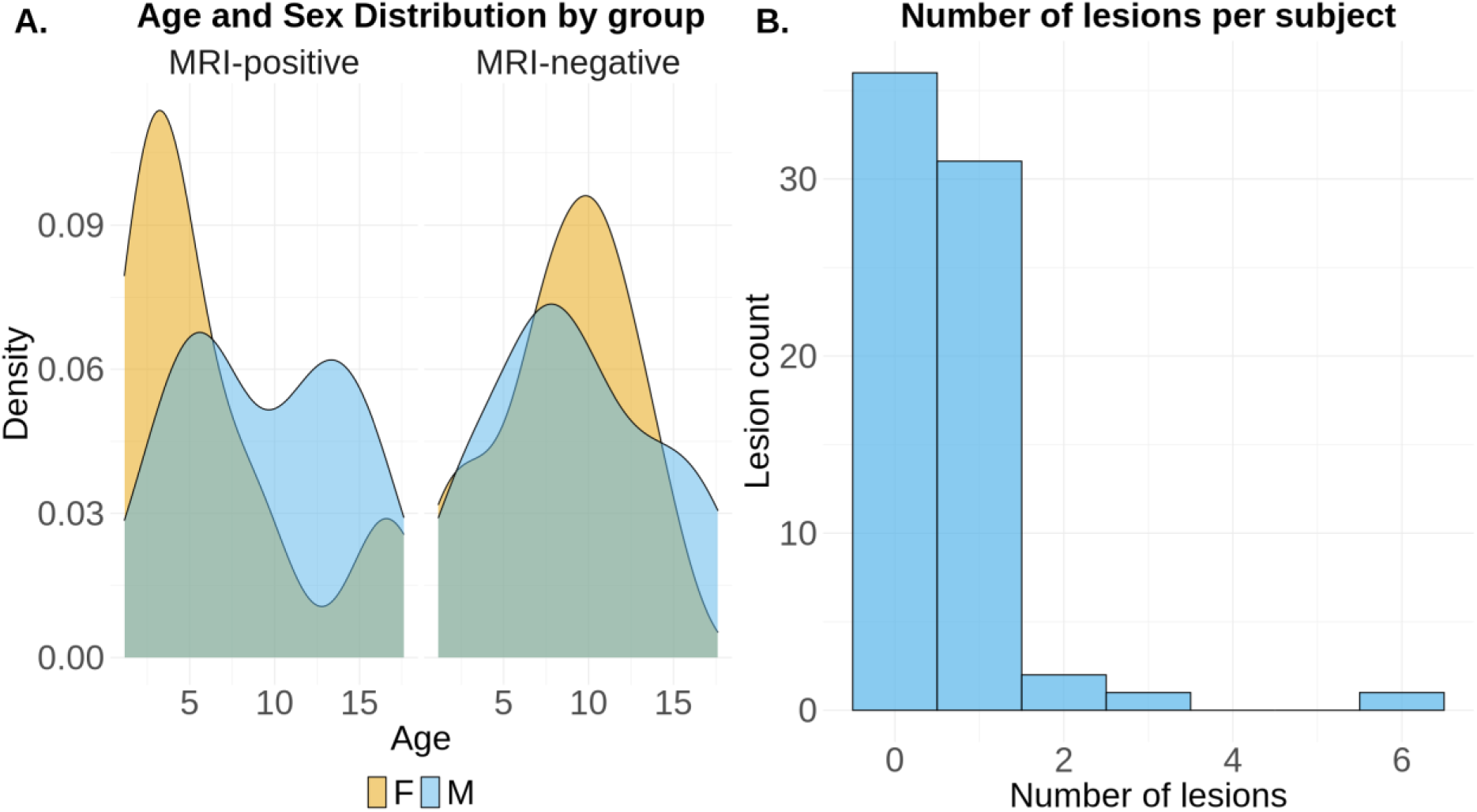
(A) Age and sex distribution (F: female, M: male) of MRI-positive (MRIpos) and MRI-negative (MRIneg) patients. B. Distribution of the number of lesions per subject across the MRIpos cohort, illustrating that most patients present with a single lesion, while a small subset exhibits multiple lesions.

**Table 2:** Demographics.

|  | MRI-POSITIVE | MRI-NEGATIVE | TOTAL |
| --- | --- | --- | --- |
| **N** | 35 | 36 | 71 |
| **AGE (YEARS, MEDIAN [IQR])** | 5.51 [3.26-11.91] | 8.78 [6.48-11.70] | 7.67 [3.66-11.71] |
| **% FEMALE** | 57.14 | 50.00 | 53.52 |

### Quality Control of FreeSurfer Segmentations

Following QC, five patient scans were rated as borderline (score 2), including three MRIpos (aged 1,2 and 2 years) and two MRIneg cases (aged 1 and 7 years old). These cases accounted for three FPLs, two FNLs, and one TPL. No exclusions were made based on QC. FSQC metrics and QC examples are reported in Supporting Fig. S1-3.

Additionally, to evaluate the potential impact of excluding these borderline cases, we additionally report lesion-based and patient-based analyses with these subjects excluded in Supporting Information Fig. S4.

### Neuroradiological and clinical re-evaluation following MELD Graph analysis

Agreement between MELD Graph and the primary report was observed in 23 lesions (TPLs), while MELD Graph identified 23 lesions that could not be verified by the neuroradiologist (FPLs). In 21 cases, lesions were detected by the neuroradiologist but not by MELD Graph (FNLs). Importantly, two rTPL were identified - lesions initially overlooked by the neuroradiologist but confirmed upon re-evaluation. Lesions classified as retrospectively true-positive (rTPL) on post hoc re-evaluation were reported descriptively but were not incorporated into the predefined reference standard and therefore were excluded from formal performance calculations. Examples for each category are given in Fig. 2.

**Figure 2:**
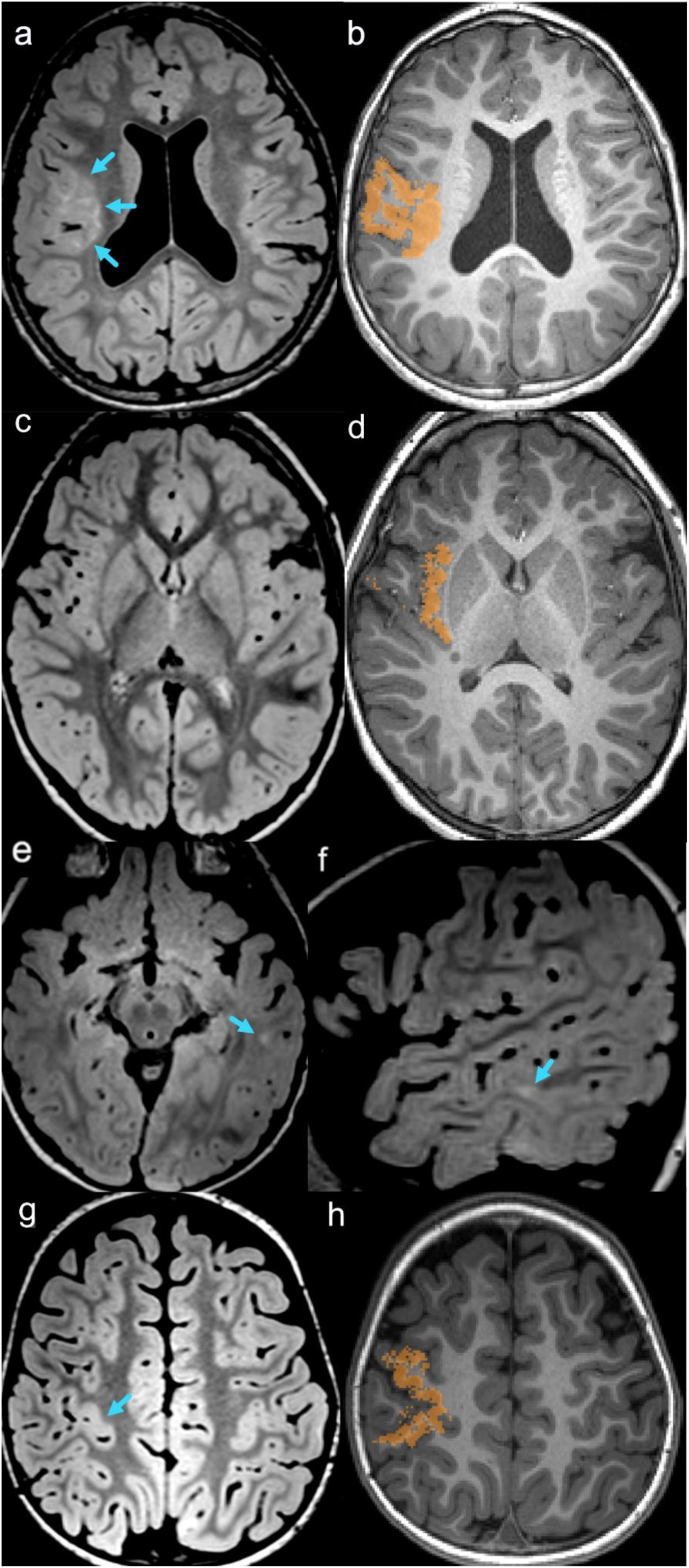
Representative imaging examples: (a–b) Correctly identified lesion (TPL) in the right insula consistent with presumed FCD type 2A, shown on FLAIR (blue arrows, a) and the corresponding MELD Graph lesion mask on 3D T1-weighted MRI (b). (c–d) False-positive MELD Graph detection (FPL) (d) with the corresponding FLAIR image (c). (e–f) Lesion missed by MELD Graph (FNL), visible on FLAIR (blue arrows, g, h). (g–h) Retrospectively identified lesion on FLAIR (rTPL) (blue arrow, e) with the corresponding MELD Graph mask on 3D T1-weighted MRI (f).

The EEG findings and clinical semiology of patients with FPL and rTPL results were re-evaluated. In one FPP case with a left frontal lesion detected by MELD Graph, the EEG demonstrated a central focus that was considered potentially concordant with the MELD Graph finding. In the remaining FPL cases, EEG and semiology were not consistent with the MELD Graph-identified lesions. Similarly, both rTPL cases showed no concordance between imaging findings and the clinical data.

Among the TPL, most were located in the frontal lobe (n = 10), followed by the temporal (n = 6), parietal (n = 4), and frontoparietal regions (n = 2). Considering both MRIpos and MRIneg patients together, the 23 FPL were found in the temporal (n = 9), frontal (n = 6), frontoparietal (n = 2) and parietal (n = 4), with single occurrences in the insula, and cingulate. FNL occurred predominantly in the frontal lobes (n = 10), followed by the temporal and parietal lobes (n = 5 and n = 3, respectively) and one case in both occipital lobe and cingulate gyrus. The two rTPL were in the frontoparietal and the frontal lobes. The overall distribution is illustrated in Fig. 3. Subsequent review of the rTPL cases by clinical and EEG evaluation did not support any of these detections as epileptogenic lesions.

**Figure 3:**
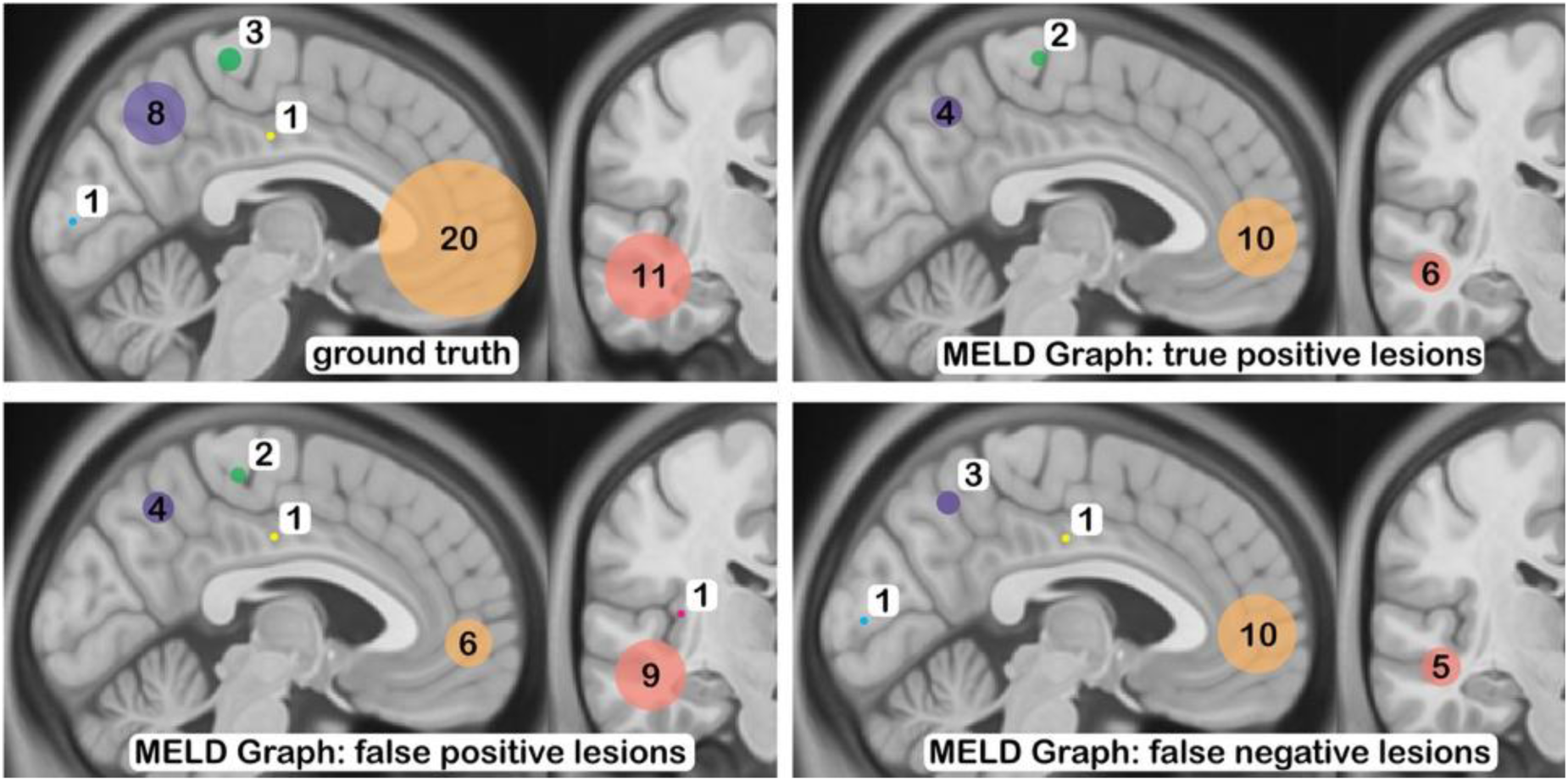
Distribution of ground-truth lesions based on the original radiology reports and the corresponding true positive, false positive, and false negative detections generated by MELD Graph, stratified by anatomical location (orange: frontal; **light red**: temporal; purple: parietal; green: frontoparietal; turquoise: occipital; yellow: cingulate, red: insula).

### Neuroradiological and clinical re-evaluation following 3D-nnUNet analysis

The model correctly identified 21 of the 44 ground truth lesions (TPL). These were in the frontal lobe (n = 10), temporal lobe (n = 5), parietal lobe (n = 3), frontoparietal region (n = 2), and occipital lobe (n = 1). In addition, the model produced 7 FPL, including four in the frontal lobe and one in the insular region, as well as two detections erroneously located outside the brain.

A total of 23 of the 44 lesions were missed by the model and classified as FNL. These were in the frontal (n = 9), parietal (n = 6), and temporal lobes (n = 6), with single lesions in the frontoparietal region and the cingulate cortex. No rTPL were identified.

Examples for each category are given in Fig. 4, and the overall distribution is shown in Fig. 5.

**Figure 4:**
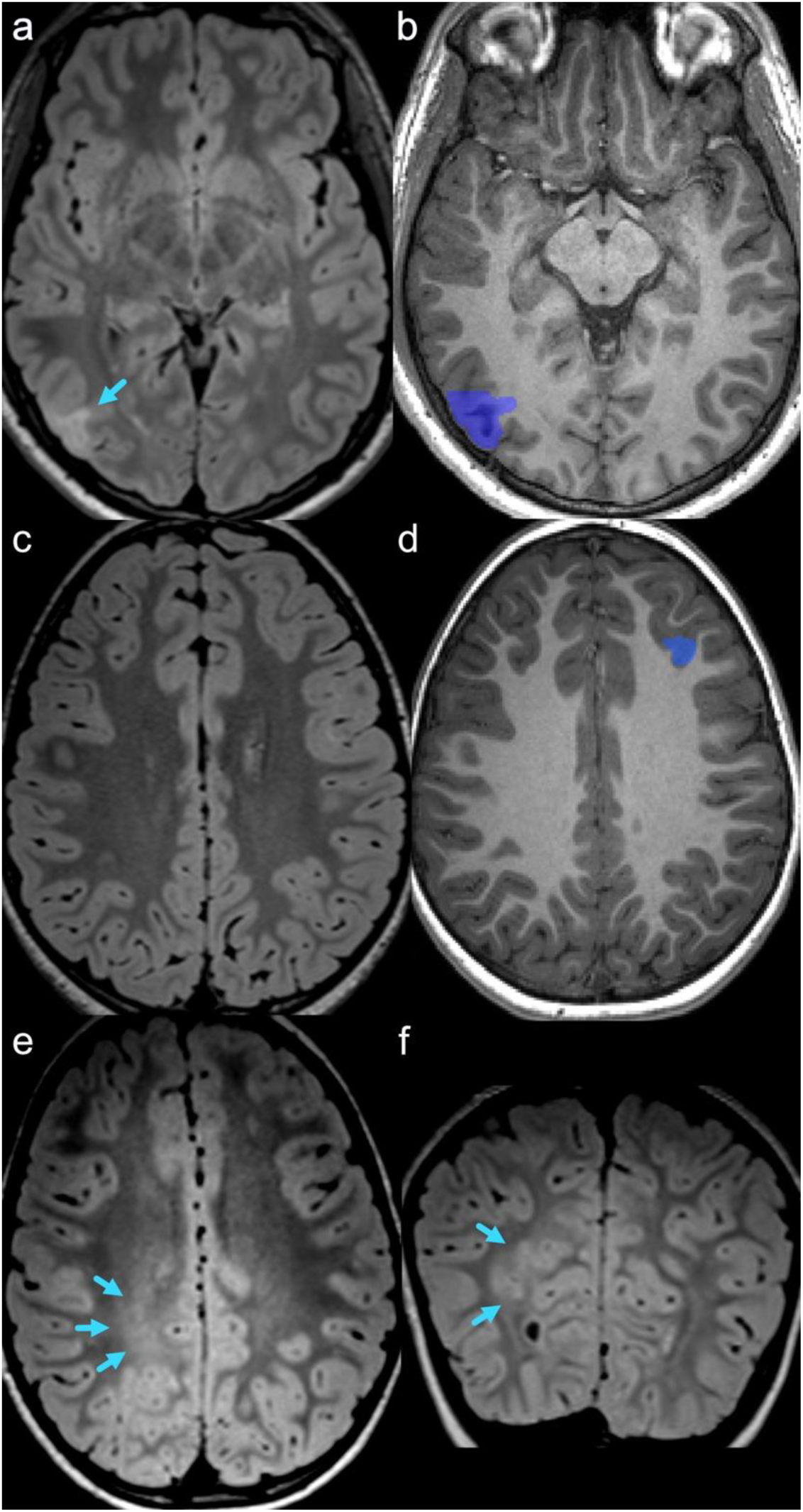
Representative imaging examples. (a–b) Correctly detected lesion (TPL) in the right temporal lobe consistent with presumed FCD type 2B, demonstrated on FLAIR (blue arrow, a) with the corresponding 3D-nnUNet segmentation mask on the 3D T1-weighted image (b). (c–d) False-positive detection (FPL) by 3D-nnUNet (d) with the corresponding FLAIR image shown in (c). (e–f) Lesion not detected (FNL) by 3D-nnUNet but visible on FLAIR (blue arrows, e, f).

**Figure 5:**
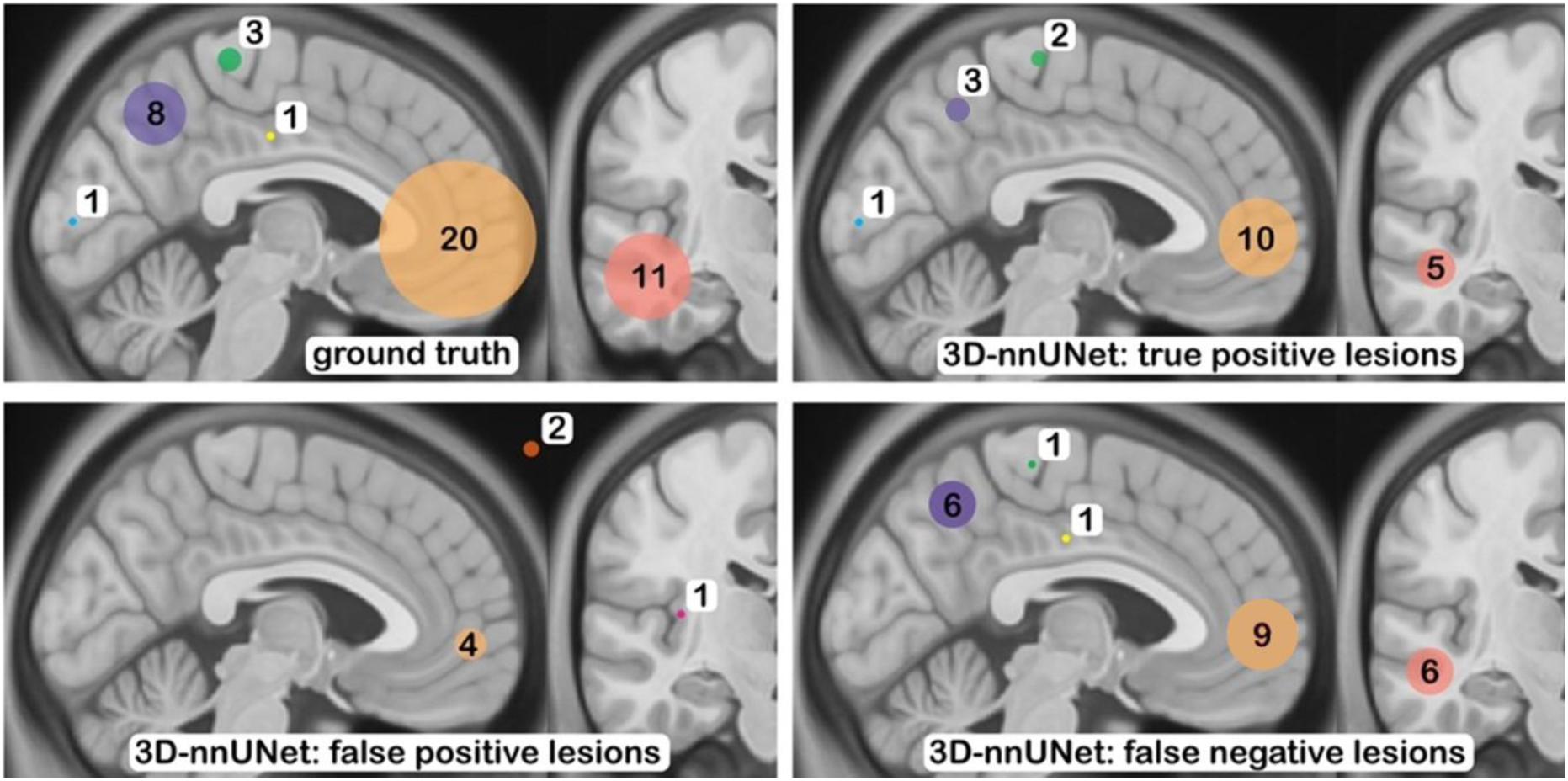
Comparison of lesion locations reported in the original radiology reports (ground truth) with corresponding 3D-nnUNet classifications as true positive, false positive, and false negative findings. Circle colors indicate anatomical region. Orange: frontal; **light red**: temporal; purple: parietal; green: frontoparietal; turquoise: occipital; yellow: cingulate, red: insula).

### Comparison between MELD Graph and 3D-nnUNet

At the lesion level, the two models showed substantial overlap, with 14 ground-truth lesions detected by both models and 14 missed by both. Among the discordant lesions, 9 were detected only by MELD Graph and 7 only by 3D-nnUNet. At the patient level, both models correctly identified 15 of 35 MRIpos patients and correctly classified 19 of 36 MRIneg patients. MELD Graph additionally identified 7 MRIpos patients missed by 3D-nnUNet, whereas 3D-nnUNet additionally identified 4 MRIpos patients missed by MELD Graph. In the MRIneg group, MELD Graph produced false-positive classifications in 12 patients classified correctly by 3D-nnUNet, whereas 3D-nnUNet produced a false-positive classification in 1 patient classified correctly by MELD Graph.

### Statistical evaluation

#### Lesion detection

In the lesion-detection task, the two models showed broadly similar performance within the MRI-positive group. MELD Graph achieved a precision of 0.85 and a recall of 0.52, whereas 3D-nnUNet achieved a precision of 0.91 and a recall of 0.48. In the MRI-negative group, MELD Graph produced 19 false-positive lesions (0.53 per patient), whereas 3D-nnUNet produced 5 false-positive lesions (0.14 per patient). MRI-negative scans without false-positive detections were observed in 20/36 (0.56) patients for MELD Graph and 31/36 (0.86) patients for 3D-nnUNet. Metrics for the lesion-detection task are shown in Fig. 6 and summarized in Table 3. Detailed counts are provided in Supporting Information Table S1.

**Figure 6:**
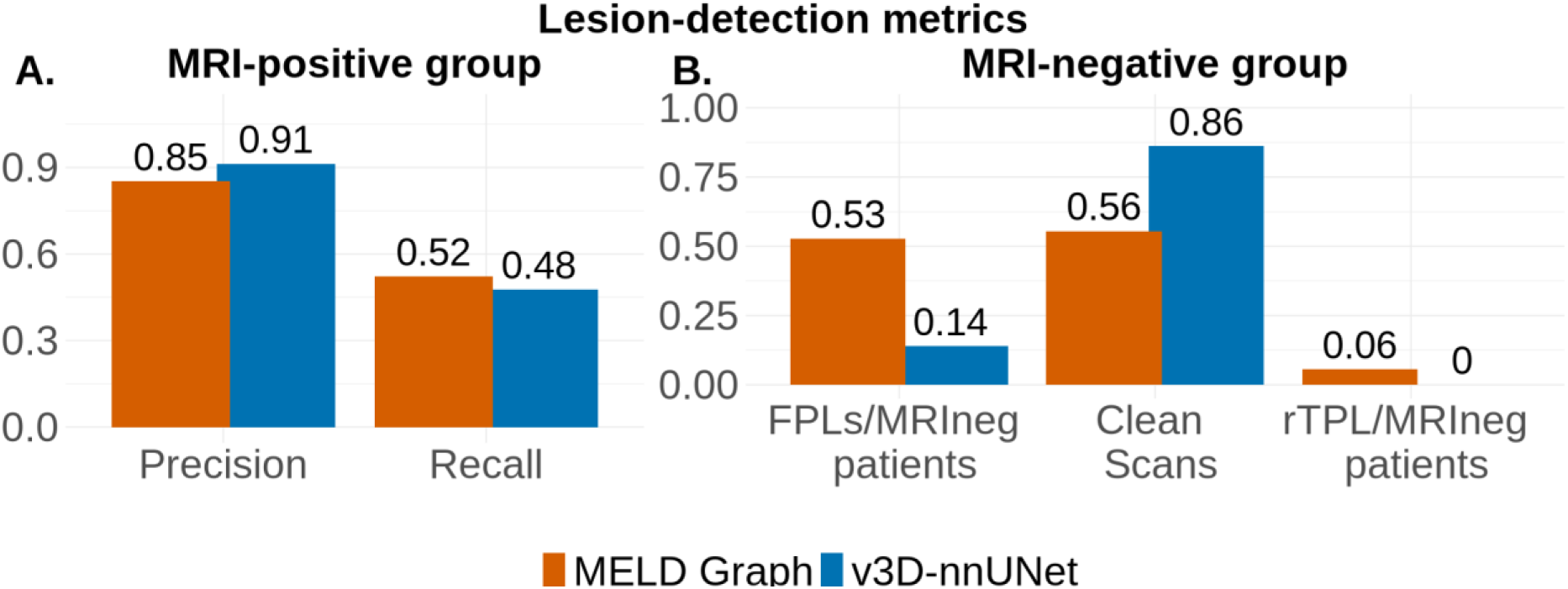
Metrics for the lesion detection task. A: In the MRI-positive group, both models show comparable performance. B: In the MRI-negative (MRIneg) group, 3D-nnUNet produces fewer false positive lesion detections (FPLs) per MRIneg patients, likely reflecting a more conservative strategy deriving from the neural network architecture and from the ensemble strategy. In contrast, MELD Graph identified additional focal cortical dysplasias (FCDs) that were not described in the primary radiology report (rTPL).

**Table 3:** Comparison of performance metrics between the present study and previous similar studies. There is to note that part of the variability across studies may also reflect differences in cohort definition. CI, confidence interval. (1) Recalculated from aggregate counts kindly provided by the original authors to align the metrics with those used in the present study. In the original paper, MRI-positive and MRI-negative cases were analyzed together.

| STUDY | THIS STUDY |  | RIPPART ET AL. [16] | KERSTING ET AL. [14] | DE VITA ET AL. [17] | KAAS ET AL. [23] |
| --- | --- | --- | --- | --- | --- | --- |
| **MODEL** | MELD Graph | 3d-nnUNet | MELD Graph | 3D-nnUNet | MELD Graph | MELD Graph |
| LESION DETECTION TASK - MRIPOS |  |  |  |  |  |  |
| **PRECISION/PPV** | 0.85 | 0.91 | 0.76 | 0.63 | 0.58 | 0.63 (1) |
| **95% CI** | 0.64-0.96 | 0.73-1.00 | 0.61-0.79 | - | - |  |
| **RECALL/SENSITIVITY** | 0.52 | 0.48 | - | 0.55 | 0.65 | 0.31(1) |
| **95% CI** | 0.35-0.66 | 0.33-0.61 | - | - | - |  |
| LESION DETECTION TASK - MRINEG |  |  |  |  |  |  |
| **FPL/PATIENT** | 0.52 | 0.14 | - | - | - | 0.66 |
| PATIENT CLASSIFICATION TASK |  |  |  |  |  |  |
| **SENSITIVITY** | 0.63 | 0.54 | 0.72 | 0.64 | - | 0.76 |
| **95% CI** | 0.45-0.78 | 0.37-0.70 | 0.61-0.78 | - | - | 0.67-0.84 |
| **SPECIFICITY** | 0.56 | 0.86 | 0.56 | 0.86 | - | 0.47 |
| **95% CI** | 0.38-0.72 | 0.73-0.87 | 0.47-0.66 | - | - | 0.38-0.56 |

#### Patient classification

MELD Graph achieved a sensitivity of 0.63 and specificity of 0.56, whereas 3D-nnUNet achieved a sensitivity of 0.54 and a specificity of 0.86 reflecting the reduced number of FPPs. Metrics for patient classification are shown in Fig. 7 and summarized in Table 3.

**Figure 7:**
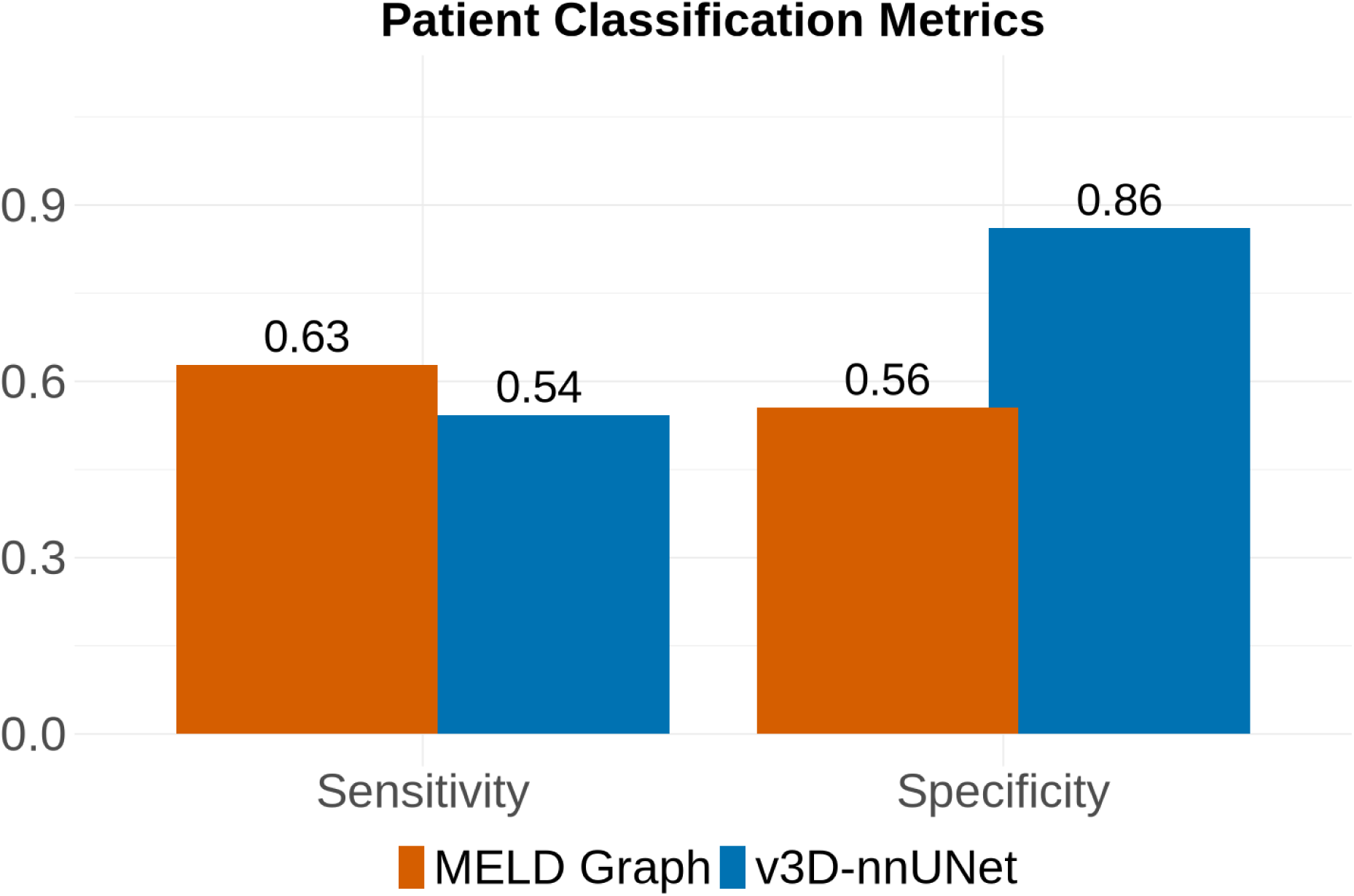
Patient-wise metrics. The two models achieved similar sensitivity values, reflecting the comparable number of correctly classified MRI-positive patients, while 3D nnUNet demonstrated higher specificity, reflecting the lower number of false-positive classifications (FPL). However, these differences did not reach statistical significance.

#### Statistical comparison

In the mixed-effects logistic regression model, no significant effect was observed for model (OR = 0.79, p = 0.58), FLAIR type (OR = 1.42, p = 0.49), age (OR = 1.01, p = 0.81) or sex (OR = 0.78, p=0.61).

For the patient-level outcome, logistic regression showed no significant association for model, FLAIR sequence, age, or sex. Correct patient classification was higher for v3D-nnUNet than for MELD Graph (OR = 1.67, p = 0.15), and higher for FLAIR250 than for the reference FLAIR sequence (OR = 2.03, p = 0.07), although neither effect reached statistical significance. Age (OR = 1.02 per year, p = 0.60) and sex (OR = 1.37, p = 0.40) were also not significantly associated with patient-level performance.

For false positives, 3D-nnUNet showed significantly lower odds than MELD Graph under the reference FLAIR condition (OR = 0.13, p = 0.002). FLAIR250 was associated with lower false-positive odds, although not significantly (OR = 0.45, p = 0.162), and the model-by-FLAIR interaction was not significant (OR = 5.79, p = 0.074). For false negatives, neither model nor FLAIR showed a significant association (OR = 0.53, p = 0.274), and the interaction was not significant (OR = 0.57, p = 0.498). The results are summarized in Fig. 8.

**Figure 8:**
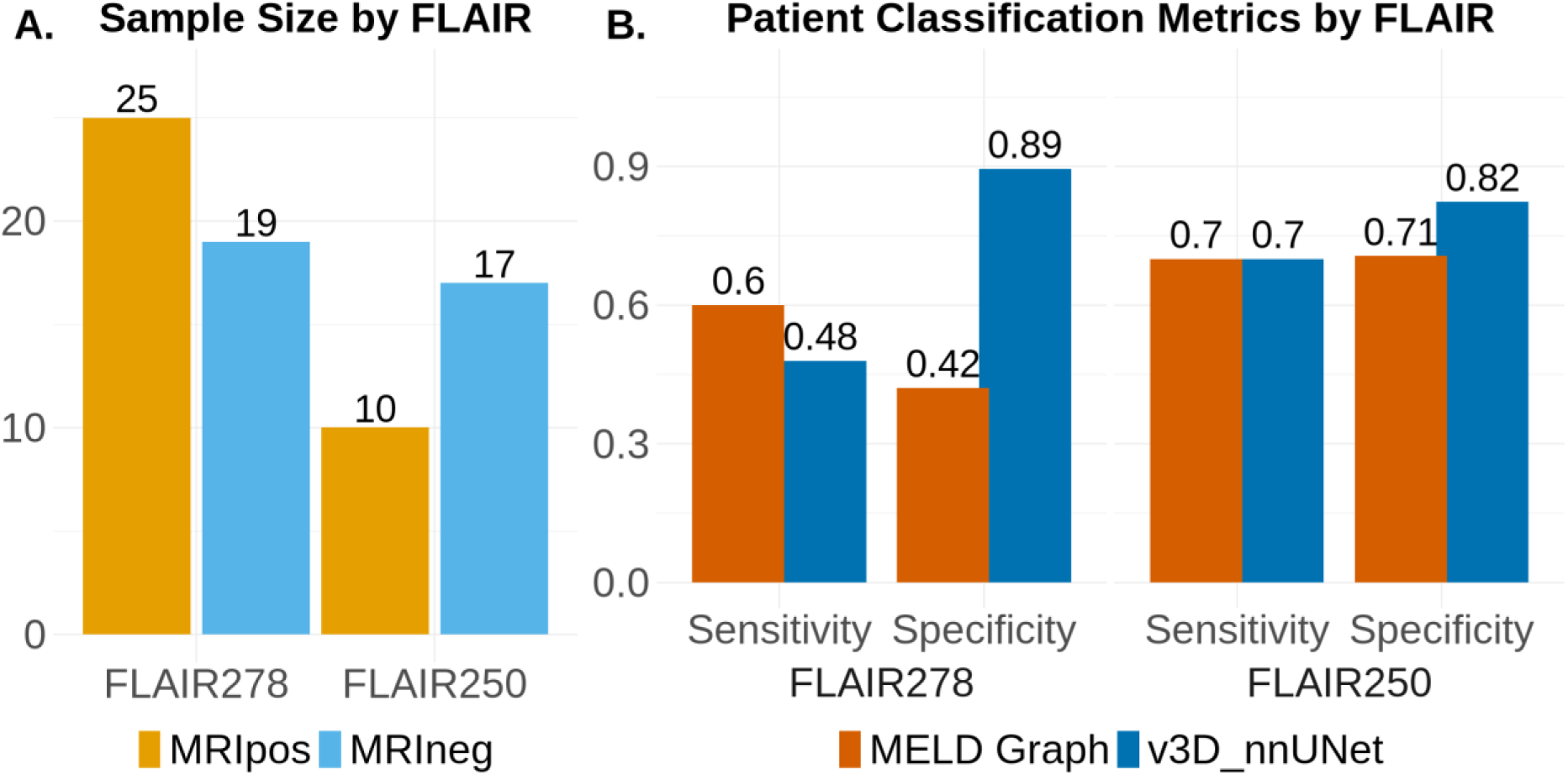
A. Sample size stratified by group and FLAIR versions. The dataset contains significantly fewer MRI-positive cases for the improved FLAIR250 sequence. B. The improved FLAIR sequence is associated with increased specificity for MELD Graph and increased sensitivity for 3D-nnUNet; however, these differences did not reach statistical significance.

## Discussion

In this retrospective study, we externally validated two recent deep-learning–based FCD lesion segmentation models, MELD Graph and 3D-nnUNet, in a cohort of children and adolescents. The central neuroradiological challenge is to identify a structural lesion that may explain the epileptogenic focus and guide appropriate therapeutic decision-making. As these lesions can be very subtle and because seizures and EEG abnormalities may also occur in structurally normal-appearing brains, we deliberately included MRIneg scans in our validation cohort.

In the lesion-detection task for the MRIpos group, the two models showed comparable performance. High precision indicates that most detected lesions represent TPL, with a small number of FPL findings in MRIpos patients. In contrast, the moderate recall demonstrates that a substantial proportion of FCDs remain undetected, meaning that neither model can reliably exclude structural pathology. Expert neuroradiological review therefore remains indispensable. In their current form, the models should be regarded as decision-support tools rather than replacements for experienced neuroradiologists.

In the MRIneg cohort, MELD Graph generated more FPL detections than 3D-nnUNet. While this “overcalling” inevitably leads to additional reassessment and increased reporting time, it may also reflect a lower detection threshold that prioritizes sensitivity over specificity. Indeed, in our cohort, two lesions initially overlooked on expert review were correctly highlighted by MELD Graph. Thus, the higher false-positive rate represents a trade-off between efficiency and sensitivity.

A potential explanation for the observed differences in false-positive occurrence lies in the fundamentally different representations used by the two models. The 3D-nnUNet operates voxel-wise, relying on local image intensity patterns. This design favors segmentation only when the learned signal is sufficiently strong, resulting in a more conservative behavior, particularly in MRIneg patients.

In contrast, MELD Graph operates on surface-derived features of the cortex. While this approach is well suited to capture cortical patterns typical of FCD, it may be more sensitive to developmental variability in the pediatric cortex. Moreover, MELD Graph relies on harmonization of surface-based features across subjects. Although our sample size exceeded the approximately 20 subjects suggested in the original publication [16], the rapidly evolving cortical morphology in pediatric populations may require larger age-stratified samples to achieve robust harmonization. In addition, FreeSurfer-based surface reconstructions are known to be less reliable in younger children and infants [27], which may further contribute to spurious surface abnormalities. MELD Graph applies the same FreeSurfer atlas across all ages. Nevertheless, our logistic regression analysis did not reveal an association between patient age and the occurrence of false positives for either method, suggesting that age alone does not fully explain the observed differences.

It should be noted that the ratio of false-positive lesions per patient was similar for 3D-nnUNet in the MRIpos and MRIneg groups. In contrast, MELD Graph produced a substantially higher number of false-positive lesions in the MRIneg group, and a similar trend can be inferred from the results reported by Kaas et al. [23]

Our findings are consistent with previous work on automated FCD detection using both voxel-wise and surface-based approaches, suggesting that both models can be generalized to a pediatric cohort. For MELD Graph, both precision and recall were broadly in line with the values reported in the original publication [16]. Compared with the original 3D-nnUNet study [14], as well as with a recent pediatric-only validation [17] and a recent mixed-cohort validation [21], our precision values were higher, whereas recall remained similar overall. An exception was Kaas et al., in which higher recall values were reported. In particular, the study by Kaas et al. included a subset of patients with structural abnormalities other than FCD, including tumors, mesial temporal sclerosis, cysts, gyral abnormalities, and infarction- or trauma-related sequelae, whereas such cases were excluded from our cohort, except for three lesions that were only recognized postoperatively as non-FCD pathologies because they could not be reliably distinguished from FCD on MRI. This difference in inclusion criteria may have influenced the pattern of false-positive detections and therefore may contribute to the discrepancies between studies.

Of note, the false-positive burden of both models was lower than that previously reported for FCD detection tools in adult and pediatric cohorts [28]. For example Gill et al [29] report an average of six false-positive cases per patient. Moreover, Kaas et al. [23] reported a substantially lower number of false-positive lesions for MELD Graph than for its predecessor, the MELD Classifier [30], or for DeepFCD [29].

In the patient-wise classification task, both MELD Graph and 3D-nnUNet achieved moderate sensitivity. This indicates that both approaches were able to correctly identify a substantial proportion of patients with FCDs, while still failing to detect a non-negligible fraction of affected patients, potentially delaying the necessary referral for epilepsy surgery. In contrast, 3D-nnUNet achieved a higher specificity compared with MELD Graph, consistent with the more conservative detection profile described above. Higher specificity may be advantageous in clinical practice, as it reduces unnecessary and time-consuming review, multidisciplinary discussions, or additional diagnostic investigations.

Two different FLAIR protocols with differing turbo factor were used in our cohort. Lower turbo factor (FLAIR250) results in reduced T2w blurring and improved signal stability, thereby enhancing grey-white matter differentiation and probably lesion border delineation. When stratifying patient-level classification performance by FLAIR image quality, we observed differential effects on the two models. For MELD Graph, higher FLAIR quality was associated with a marked increase in specificity, driven by a reduction of false positives, whereas sensitivity remained largely stable. In contrast, 3D-nnUNet showed higher sensitivity with the higher-quality FLAIR250 with specificity showing a more modest change. These findings indicate that FLAIR quality influences patient-level classification in a model-dependent manner. A plausible explanation lies in the different model representations. Because MELD Graph relies on surface-based features, a higher signal-to-noise ratio (SNR) may amplify subtle surface irregularities, which can be misinterpreted as lesion-like patterns, thereby generating false positive classifications. In contrast, the 3D-nnUNet operates voxel-wise, and from a signal-detection perspective, higher SNR can be interpreted as a reduction in background noise and a consequent increase in lesion-to-background contrast, such that lesion-induced deviations exert a larger relative effect. As a result, subtle FCD-related abnormalities are less likely to be absorbed into background variability and more likely to be detected, even after resampling to the T1w space.

However, this analysis should be interpreted cautiously, as the two FLAIR sequence groups were unbalanced in size, particularly with fewer MRIpos cases in the FLAIR250 group. Therefore, the observed differences may partly reflect differences in the underlying patient samples rather than sequence effects alone. Moreover, the limited sample size reduced statistical power, and our regression models did not detect a significant effect of FLAIR sequence.

Although the primary radiology reports and the subsequent review by an expert pediatric neuroradiologist served as the reference standard, this approach has inherent limitations. FCDs can be highly subtle and difficult to distinguish from morphologically similar pathologies. In our cohort, four lesions initially presumed to represent FCD on presurgical MRI were later histologically confirmed as gangliogliomas or gliotic tissue. Furthermore, many of the lesions classified as FCD were not confirmed histologically because biopsy was not taken. Consequently, some FNL classifications may also reflect limitations of the human reference standard rather than true model failure.

Furthermore, the reevaluation of FPL detections remains subjective. In principle, both model architectures may identify abnormalities that are not readily appreciable on visual inspection. Two rTPL were identified only after careful re-examination of all available sequences. Notably, neither correlated with the clinical findings, raising the possibility that model prompting may have influenced expert interpretation. This phenomenon is not different to a setting involving multiple experts and complex diagnostic tasks, where a degree of subjectivity is unavoidable and must be acknowledged. This finding highlights at the same time the possibility that AI-based decision-support tools may influence human interpretation by drawing attention to subtle lesion-like abnormalities; in such cases, clinical and electroencephalographic correlation is important to reduce the risk of overinterpreting findings prompted by the model.

Another limitation affecting both models and the human reader relates to variability in lesion size and spatial extent. In many cases, the delineated lesion volumes differed substantially between raters and algorithms. Minor discrepancies in lesion location were also observed, a phenomenon previously reported in the original MELD Graph publication [16]. Given this inherent variability in spatial definition, formal validation metrics assessing spatial overlap or volumetric differences were not applied. This consideration is particularly relevant when applying both automated models and human assessments to guide the extent of surgical resection. Future studies directly comparing presurgical lesion delineation, actual resection margins, and postoperative seizure outcomes are therefore essential.

Finally, this study is limited by the relatively small sample size. This can be important especially in MELD Graph where the feature harmonization step can be crucial for the occurrence of FPL. It remains uncertain whether including a larger number of subjects across different pediatric age groups, would improve the robustness of the harmonization and, consequently, model performance.

In conclusion, our findings support the generalizability of both MELD Graph and 3D-nnUNet to an independent pediatric cohort. Both models demostrated high precision in detecting FCDs within our pediatric cohort, but only moderate sensitivity, leaving a clinically relevant proportion of lesions undetected. Both models therefore represent valuable decision-support tools that may enhance diagnostic confidence, yet they are not suitable as independent screening systems. Importantly, image quality, particularly FLAIR sequence quality, may influence patient-level sensitivity and specificity, supporting the use of optimized acquisition protocols despite longer scan times.

## Acknowledgements

The authors acknowledge the Scientific Computing of the IT Division at the Charité - Universitätsmedizin Berlin and of the Berlin Institute of Health Center of Digital Health for providing computational resources that have contributed to the research results reported in this paper.

The authors also thank Helene Kaas (Neurobiology Research Unit, Copenhagen University Hospital Rigshospitalet, Copenhagen, Denmark) for kindly sharing aggregated lesion-level count data from her study, which enabled recalculation of performance metrics for comparison with the present work.

## Data Availability Statement

The raw MRI data analyzed during the current study are not publicly available due to German data protection regulations but are available from the corresponding author upon reasonable request. Defaced example FLAIR images illustrating the two acquisition variants used in this study are publicly available in the OSF repository https://osf.io/ag78q

## Author contributions statement

- Andrea Dell’Orco curated the dataset, conceived and performed the statistical analyses, interpreted the results, and drafted the manuscript
- Enrico De Vita supported the implementation of the analysis and revised the manuscript for intellectual content
- Felice D’Arco supported the implementation of the analysis and revised the manuscript for intellectual content
- Annalena Lange revised the manuscript for intellectual content
- Theodor Rüber revised the manuscript for intellectual content
- Angela M. Kaindl contributed patients, participated in the discussion of the clinical, electrophysiological, and radiological data, and critically reviewed the manuscript.
- Mike Wattjes contributed to the collection of radiological data and critically reviewed the manuscript.
- Ulrich-Wilhelm Thomale contributed patients, radiological data and critically reviewed the manuscript.
- Lena-Luise Becker contributed patients, participated in the discussion of the clinical, electrophysiological, and radiological data, and critically reviewed the manuscript.
- Anna Tietze conceived and supervised the study, curated the dataset, contributed to data acquisition and clinical review, contributed to the interpretation of the findings and drafted the manuscript.

