## Supporting Information for "Deep Learning-Based Detection of Focal Cortical Dysplasia in Children: External Validation of the MELD Graph and 3D-nnUNet pipelines"

### S.1. Formulas

$$Precision = \frac{TPL}{(TPL + FPL)}$$

$$Recall = \frac{TPL}{(TPL + FNL)}$$

$$Sensivity = \frac{TPP}{(TPP + FNP)}$$

$$Specificity = \frac{TNP}{(TNP + FPP)}$$

### Tables

*Table S1: Summary of lesion-level and patient-level classification counts and performance metrics for MELD Graph and 3D-nnUNet*

| Task | Metric | MELD Graph | 3D-nnUNet |
|---|---|---|---|
| Lesion detection (MRI-positive patients) | True positive lesions (TPL) | 23 | 21 |
| | False positive lesions (FPL) | 4 | 2 |
| | False negative lesions (FNL) | 21 | 23 |
| | Retrospectively true positive lesions (rTPL) | 2 | 0 |
| | Precision | 0.85 | 0.91 |
| | Recall | 0.52 | 0.48 |
| Lesion detection (MRI-negative patients) | False positive lesions (total) | 19 | 5 |
| Patient classification (all patients) | True positive patients (TPP) | 22 | 19 |

| | | | |
|---|---|---|---|
| | False positive patients (FPP) | 13 | 16 |
| | False negative patients (FNP) | 16 | 5 |
| | True negative patients (TNP) | 20 | 31 |
| | Sensitivity | 0.63 | 0.54 |
| | Specificity | 0.56 | 0.86 |

### Figures

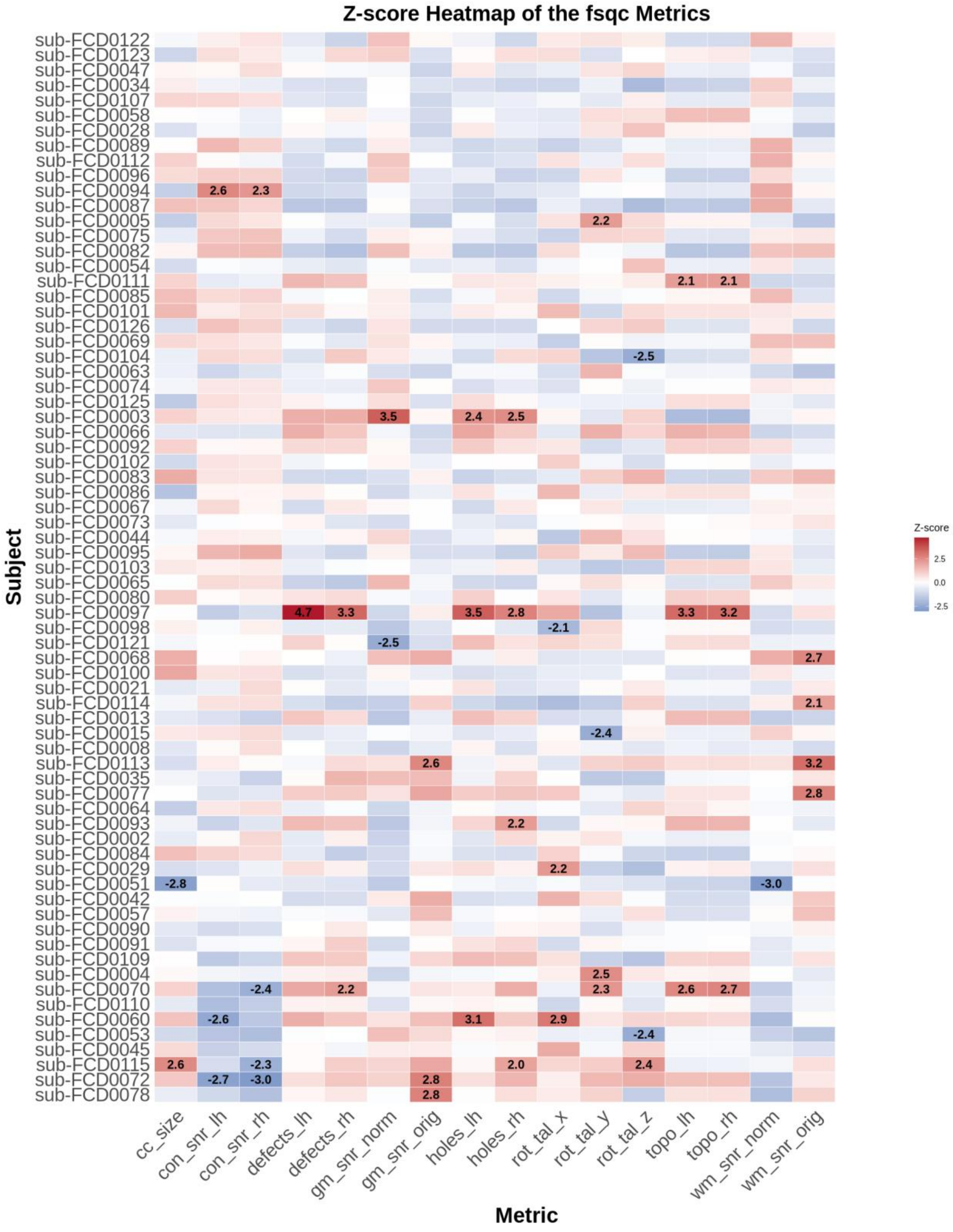


*Figure S1: Heatmap of the FSQC output metrics. Patients are ordered by decreasing age. Visual quality control resulted in a borderline rating (QC = 2) for sub-FCD0053, sub-FCD0060, sub-FCD0072, sub-FCD0097, and sub-FCD0110.*

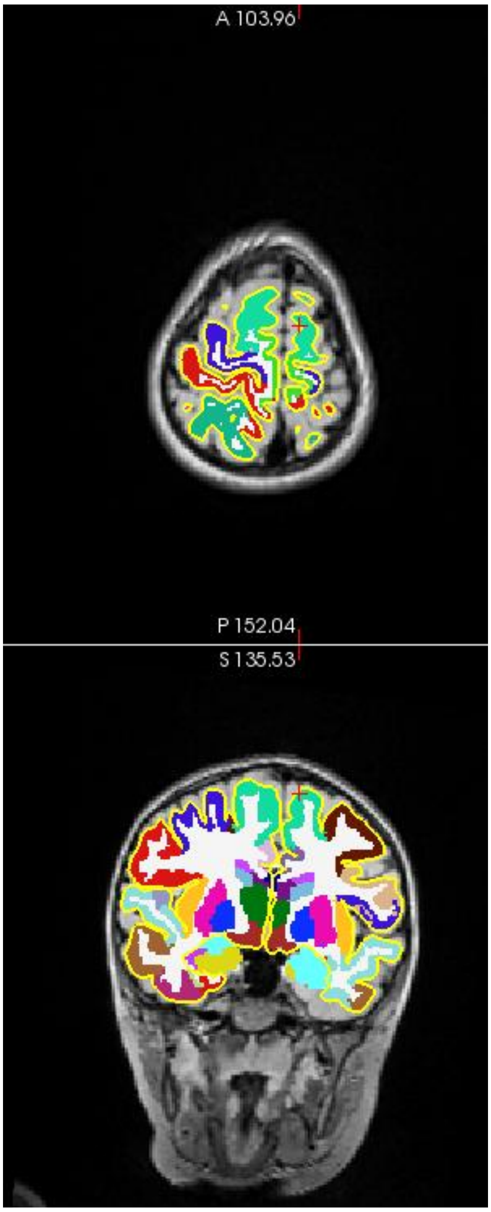


*Figure S2: Example of a borderline case. The cortex is slightly undersegmented, while the labels remain largely symmetrical.*

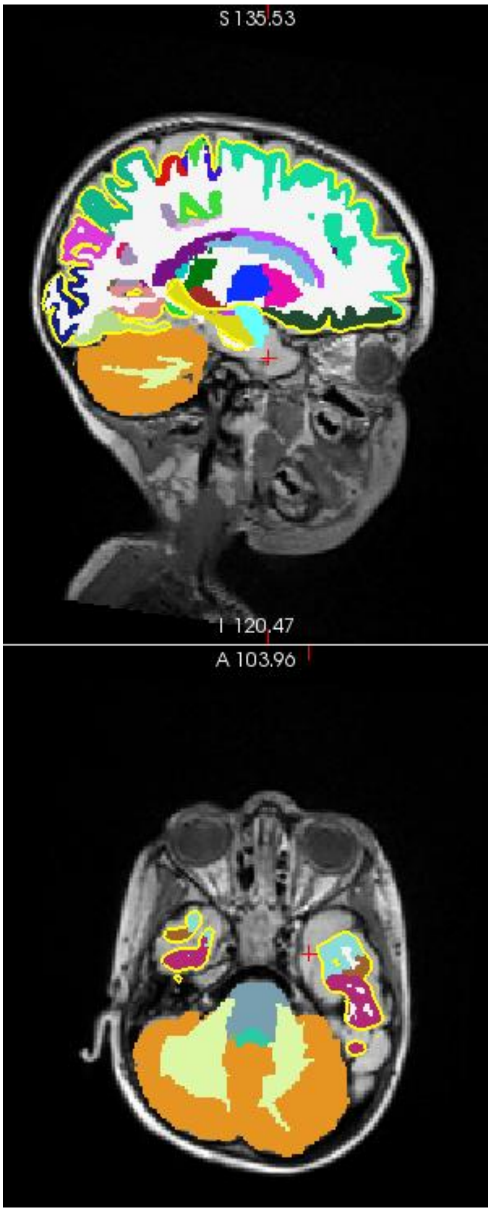


*Figure S3: Example from the visual quality control. The large temporal lobe FCD is not assigned to a segmentation ROI, which likely contributed to outlier FSQC values. However, the overall segmentation and cortical reconstruction are correct. The lesion was correctly detected by both MELD Graph and 3D-nnUNet.*

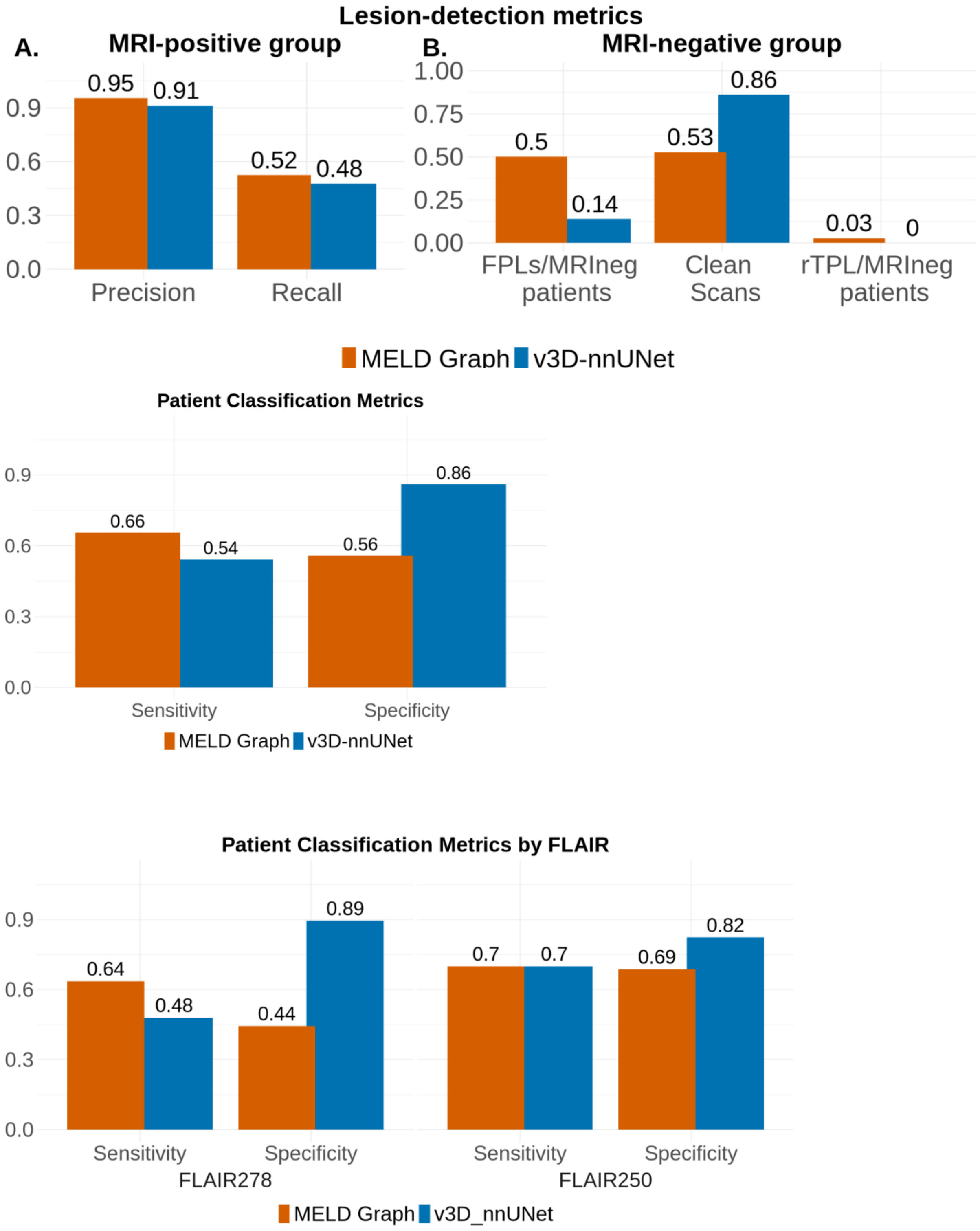


*Figure S4. Performance metrics recalculated for MELD Graph after excluding cases classified as borderline by the FreeSurfer quality control. Metrics improved slightly, indicating that some borderline cases contributed to false-positive or false-negative classifications. However, these cases were retained in the main analysis because the quality control did not support their exclusion and the overall performance changed only marginally.*
